# Specifying prospective compartmental models of chronic disease

**DOI:** 10.64898/2026.01.14.26344075

**Authors:** Tim Wilson, Samantha Howe, Stephanie Khuu, Abraham Flaxman, Tony Blakely

## Abstract

**Background:** Determining the impact of preventive interventions on future health and economic outcomes in a given population often requires parameterising disease models with forecast disease incidence, remission and case fatality rates. This paper outlines a method to specify these forecasts, using period estimates over several years from source data that are not necessarily coherent from a cohort perspective.

**Methods:** For a given chronic disease, the model is specified by first obtaining and smoothing source data inputs. We used Global Burden of Disease (GBD) period estimates of disease prevalence (*p*) incidence rates (*i*) and mortality rates (*d*) from 1990 to 2019 as a starting point, from which case fatality rates (*f*) were calculated. Values were smoothed by birth cohort within each calendar year using a quadratic Bezier curve, from which remission rates (*r*) were inferred. Next, each measure was projected for 20 years using log-linear regression by year, with 10-year age knots, produced independently by sex. Finally, a stochastic optimization algorithm was used to adjust *i,f* and *r* to ensure longitudinal (cohort)-coherence such that they matched simulated forecasts of *p* and *d*, which were treated as the reference targets.

**Model output:** The method is illustrated by presenting model output for the 20 highest disability-adjusted life year-causing chronic diseases in Australia. Model fit was assessed visually, as well as through the relative and absolute differences between calibration output and targets (*p* and *d*). A subset model using GBD input data from 1990-2009 only was also produced and compared to 2019 GBD data to investigate the model’s predictive validity.

**Conclusions:** Our method is a useful optimization procedure to derive cohort-coherent estimates of chronic disease in simulation models. For this analysis, readily available GBD period rates were used as input data; however, other raw data sources could also be applied for a given disease.

## Background

### Motivation

Priority setting and decision-making in health policy are supported by estimates of the health burden from different diseases. This burden can be quantified in different ways, for example:

1. Quantifying total burden in a year or over time, e.g., quantifying the morbidity and mortality caused by cardiovascular disease from 1990-2021. (1) This describes the size of the problem.
2. Attributing disease burden to risk factors, e.g., quantifying the cardiovascular disease burden attributed to past tobacco use vs. salt intake helps direct prevention efforts. (2)
3. Determining how much future disease burden could be avoided (or health gain achieved) if a preventive, screening, or treatment intervention were implemented (e.g. (3)).

Existing burden of disease analyses such as those produced by the Global Burden of Disease (GBD) study (4) and others (e.g., the Australian Burden of Disease Study (5)) address the first two points above. However, the third point requires forward-looking analyses of disease burden that could be avoided under intervention scenarios, something that is not routinely done in burden of disease analyses. Estimating avoidable burden can be achieved using computational modelling to simulate future population outcomes under different intervention conditions affecting disease morbidity and mortality. (6) But before simulating interventions, business-as-usual forecasts of disease rates are required.

### The problem

Compartmental models are often used to represent the way individuals or cohorts move between health states over time and are a standard approach for estimating the effect of health interventions on disease outcomes. (7) In these models, a population is divided into compartments that represent mutually exclusive states, for example moving from susceptible to infected to recovered in the classic SIR model for infectious diseases. (8) For chronic and non-communicable diseases, the same framework can be applied using compartments that estimate how many people are at risk (susceptible), how many are living with the disease (prevalent), and how many die at each timestep. (9) Movement between compartments is governed by rates of incidence, remission and fatality. Such a system is solved iteratively and at each time step the prevalence of a given outcome in the next year is determined by incidence, fatality, and remission rates in the current and past years (as shown in Figure 1).

**Figure 1.**
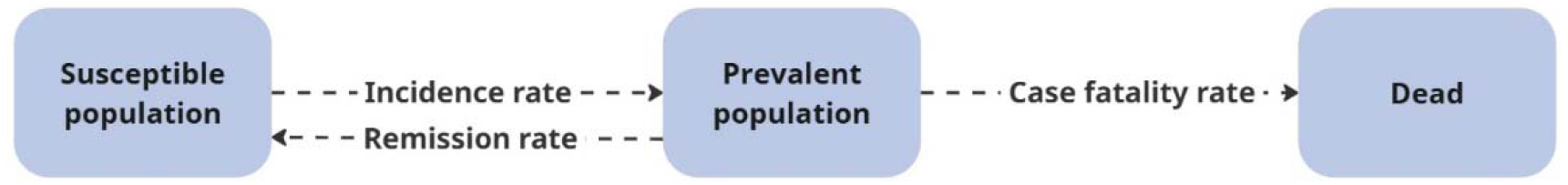
Compartmental disease model, where the prevalent population in each year is determined by incidence, remission, and case fatality rates for a disease of interest, the susceptible population is those alive who do not have the given disease, and ‘dead’ is those who have died from the disease, in a given period.

There are two challenges to work through to produce such a compartmental disease model:

1. Limited disease rate data to use for forecasting: Disease prevalence is easier to measure (e.g., through a survey) and therefore more readily available than disease incidence, case fatality, and remission rate data (hereafter referred to as ‘disease rate’ data). However, it is the disease rates that are directly altered by an intervention. Preventive interventions primarily modify the incidence rate, while treatments alter one or both of case fatality and remission rates (and potentially disease severity, which is beyond the scope of this paper).
2. Lack of coherence in disease rates: Historical disease data is generally imperfect due to factors including but not limited to measurement error, missing data, and population changes. As a result, the available disease rates (either sourced directly or inferred) are unlikely to be cohort-coherent over time. Put another way, as a simulated cohort ages through the available age-specific disease rates, implausible future prevalence and mortality rates may arise. Initial and forecast disease rates must be adjusted to ensure internal consistency across cohorts and over time.

There is no single gold-standard approach for optimizing compartmental disease models. In practice, researchers apply a broad toolkit based on available data, computational resource, and research aim. Domain-specific methodologies include those developed by the Global Burden of Disease (GBD) study team, including DisMod (10, 11), which uses Bayesian meta-regression, and CODEm (12), which uses ensemble modelling. Both approaches are largely driven by the first methodological challenge outlined above, aiming to produce estimates for all diseases and countries from sparse data to support historic burden calculation. These models provide valuable reference frameworks but can be computationally intensive when applied at scale and are not specifically built to support prospective simulation modelling.

Other optimization search algorithms have also been applied to specific disease model calibration. For example, both simulated annealing and genetic algorithm approaches have been used to calibrate the Lung Cancer Policy Model that simulates lung cancer progression. (13) These global optimization methods can be more efficient compared to regression-based or manual calibration, particularly when there are many parameters to be calibrated simultaneously.

Our proposed method draws on existing optimization approaches and presents a novel chronic disease rate generator. The aim of this method is to fill a gap in chronic disease modelling methodologies by turning limited historic disease data into coherent future disease rates, specifically suited for prospective modelling of multiple diseases. Standalone code for the generator is available at: github.com/population-interventions/CompartmentalDiseaseGenerator.

In this paper, we present the model calibration process and outputs for the top 20 causes of disability-adjusted life years (DALYs) in Australia, using GBD data (14) as the input. The rate generator can be applied to other disease datasets that have the appropriate raw data.

## Methods

### The algorithm

Our chronic disease model is based on a standard compartmental framework that tracks transitions between the susceptible and prevalent populations:

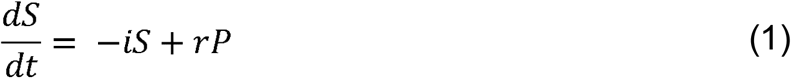

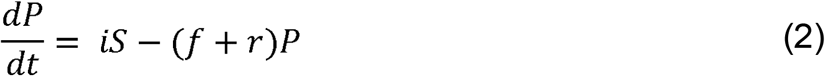

where *S* and *P* are susceptible and prevalent populations, and *i, f*, and *r* are incidence, case fatality, and remission rates, respectively. In the full model, these rates vary by age, sex, and year, so we only use equations 1-2 to calculate prevalence and mortality within a single year, within which *i, f*, and *r* are assumed constant.

The chronic disease rate generator produces the required parameters via a three-step process, shown in Figure 2 and Box 1. Step 1 smooths the input data across age and birth cohorts and infers the underlying remission rate that is not present in the data. In step 2 regression models estimate annual rate changes, resulting in two groups of forecasts:

**Figure 2:**
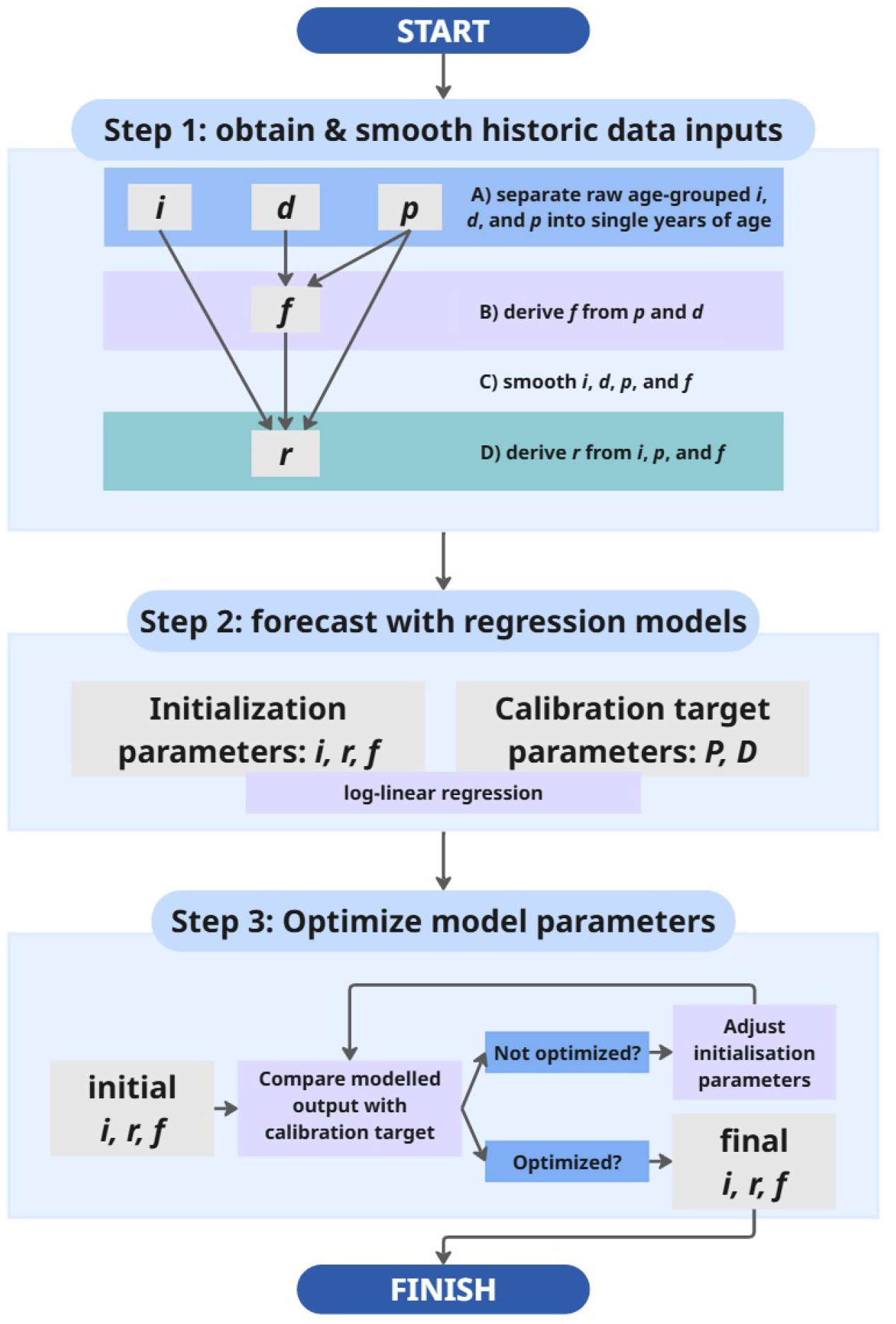
Algorithm conceptual diagram, where *p*= prevalence risk, *d*= mortality rate, *i*= incidence rate, *f*= case fatality rate, *r*= remission rate, *P*= prevalence (count), *D*= death (count).

- Calibration target parameters: prevalence, mortality rate. These parameters are assumed to be more accurate than the disease rate values and are therefore treated as the ‘ground truth’ for the following calibration step.
- Initial rate parameters: incidence, remission, and case fatality rates

Step 3 involves a stochastic optimization algorithm that iteratively adjusts the rate parameters, so that the resulting simulated prevalence and mortality rates align with the directly forecast calibration targets, therefore producing a coherent compartmental model.

The variables defined in each step are listed in Table 1. Additional detail for calculations in each step is provided in the following sections.

#### Box 1 Methods summary

**Step 1: obtain and smooth historic data inputs**

This occurs over 4 sub-steps:

A. Obtain raw (five-year age grouped) incidence rate (*i*), mortality rate (*d*), and prevalence (*p*) data from the GBD: within each calendar year and sex, take the 5-year rolling average of categorical age data, to produce single year of age data (from 0-99)
B. Case fatality rate (*f*) is calculated after smoothing step A, as 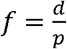
C. For *i, d, p*, and *f*: for each annual birth cohort, use a quadratic Bezier curve within each year to make a continuous function to interpolate the rate from the start to the end of each year
D. Remission (*r*) is calculated from smoothed *i, d, p*, and *f*

**Step 2: forecast for 20 years with regression models**

For *i, d, p, f*, and *r*, log-linear regression models by year, with 10-year age knots, are produced independently by sex. The resulting forecasts are split into two groups:

- Initialization parameters (*i,f,r*): the parameters that form the chronic disease model, which will be optimized in Step 3. Each initialization parameter has two sets of values: a base year value, and annual percentage change value.
- Calibration target parameters (*P, D*): yearly prevalent cases and death counts; the ‘ground truth’ parameters, used as targets for the initialization parameters to fit to in Step 3

**Step 3: optimize model parameters**

The stochastic optimization algorithm scores the modelled output using the initialization parameters against the calibration target parameters – poorly fitting parameters are adjusted, then re-scored. This occurs iteratively until a steady state is reached.

### Step 1: obtain and smooth historic disease parameters

This process was applied to historic disease-specific prevalence, mortality rate, and incidence rate data from the GBD (https://vizhub.healthdata.org/gbd-results/ (14)), specifically from the 2021 GBD release, using only data from 1991 to 2019 inclusive. Data from 2020 and 2021 were excluded, due to the impact of the COVID-19 pandemic on the diagnosis and reporting of many chronic conditions – it was assumed that these years do not reflect long-term trends. At the time of writing the 2023 GBD iteration was not publicly available for download. Other data sources could be used as the inputs for this step, or alternatively it can be skipped all together if a user has their own existing projected disease parameters.

We first apply a five-year rolling average on age to smooth discontinuities, producing single-year-of-age estimates (0-99 years) for each calendar year and sex. Sub-year smoothing is then performed by fitting the data within each year with a quadratic Bezier curve, *b*_*y*_. This is required for the inference of historic remission. Specifically:

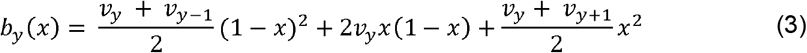

where *v*_*y*_ is the GBD value for the year-of-birth cohort in year *y*, and *x* ∈ [0, 1] ranges from the start to the end of the year. A quadratic Bezier was chosen to make the interpolation first-order differentiable across subsequent years. The derivative is given by,

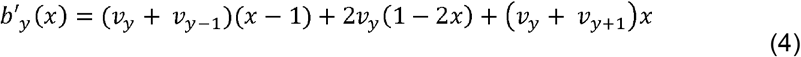

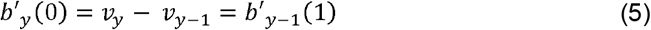

The historic data for each input are interpolated with a 0.1-year time step to produce sub-year time series for all diseases parameters.

Having created smooth sub-year curves, remission (which is not directly observed in the raw data) is inferred as follows. Let *p*(*t*), *i*(*t*) and *f*(*t*) be the Bezier curve interpolations of prevalence, incidence rate, and case fatality rate, for a fixed year-of-birth cohort, where *t* is continuous year. For example, *p*(*t*) = *b*_⌊*t*⌋_ (*t* − ⌊*t*⌋) where *b* is the Bezier curve for the prevalence of the cohort in year ⌊*t*⌋, the floor of *t*. We want to find *r*(*t*), the remission rate, by year for the cohort.

We use the following discrete approximation of the chronic disease model defined in equations 1-2, which allows the parameters to vary over time,

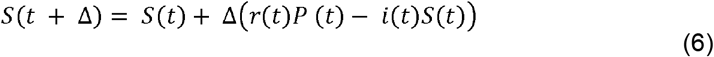

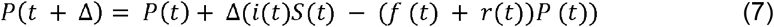

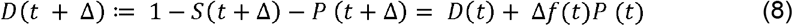

where Δ is a 0.1 timestep (as for the other parameters), is cumulative deaths *p*(*t*), is prevalent cases, and *S* (*s*) + *P* (*s*) = 1 at the start time *s*. Note that in general *P*(*t*) ≠ *p*(*t*) because the denominator of prevalence in the GBD is the current living population, so a comparison of *P* and *p* will involve scaling by 1/ (1 − *D*(*t*)).

Consider the start of the model, we know *D*(*s*) = 0, which lets us set *P*(*s*) = *p*(*s*) and *S*(*s*) = 1 − *p*(*s*). Then, considering a future timestep *s* + Δ, for the model to agree with the GBT data,

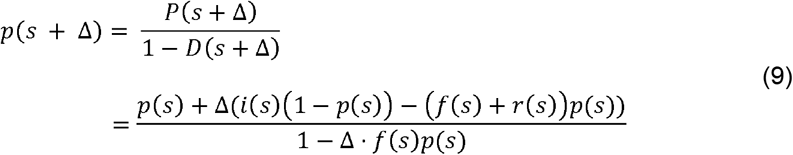

Therefore, since all the other parameters are known,

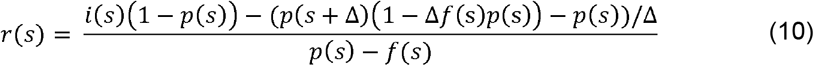

We produce *r*(*t*) for arbitrary *t* by changing the start time and repeating the above procedure. The result is smooth time series for all parameters (*i, r, p, d*, and *f*) that are internally consistent within each cohort over time.

### Step 2: projecting disease parameters

Annual percentage changes for each parameter into the future are derived via log-linear regression. The regression has age splines spaced 10 years apart from 0 to 90, and is calculated as,

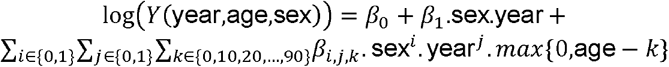

The result of the regression is only used to generate initial estimates of the annual percentage change. The initial estimate of the value in the base year is taken directly from the GBD data when available. If the selected base year falls beyond the range of available GBD data, it is extrapolated from the last observed year of data using the estimated annual percentage change.

Remission exhibits higher variability due to its dependence on other parameters. To reduce the influence of residual spikes, the remission regression is inverse square weighted by the proportional distance to the median data point. This gives greater weight to data near the centre of the age distribution and down-weights outliers. The values for both the base year remission rate and its annual percentage change are taken from the log-linear regression.

### Step 3: producing a coherent model

The initialization parameters produced by Step 2, specifically base year and annual percentage change values for incidence, case fatality, and remission rates, can be run through the full disease compartmental model to produce an estimate of prevalence and mortality. These estimates do not, in general, match the directly forecast calibration targets for prevalence and mortality. We rectify this by applying an optimization model, which iteratively adjusts the initialization rate parameters until they generate the calibration targets.

Table 2 shows the components of the calibration.

**Table 2.** Calibration components.

| Calibration component | Parameterization |
| --- | --- |
| **Variable parameters** | Base year incidence, remission, and case fatality rates<br>Annual percentage change for incidence, remission, and case fatality rates |
| **Calibration targets** | Prevalence and death count (projected from regression – Step 2) |
| **Goodness of fit (GOF) measure** | Least squares (WHO-age-weighted (15)) over 20-year horizon for both prevalence and death count |
| **Calibration update rule** | Deterministic, diminishing-step adjustments |
| **Acceptance criteria** | Parameters minimise the GOF measure |
| **Stopping rule** | Previous 20 iterations produce scores from the GOF within 0.5% of each other. |

Components based on Vanni et al. (16) checklist WHO: World Health Organisation

Let *i*_*a*_, *f*_*a*_, and *r*_*a*_ denote their annual percentage changes produced in Step 2. Let *i*_*b*_, *f*_*b*_, and *r*_*b*_, denote the initial base-year values of incidence, case fatality, and remission rates of the model. If the base year is within the GBD data window, then these are taken from the curves calculated in Step 1, otherwise they are extrapolated from the last data point using the annual percentage changes. Each of these parameters varies by age and sex; however, the model is calibrated independently for each sex. To assess how well a set of parameters fits, we initialize the model with prevalence in the base year, then simulate a future trajectory using the derived rates and their annual percentage changes and compare it to the calibration target.

To run the model, we start with our idealised compartmental model (equations 1-2). Note that *i,f*, and *r* are static in these equations, but we need them to vary over time by their annual percentage changes. Additionally, cohorts become older over time, so any given cohort should experience the incidence, case fatality, and remissions rates pertaining to increasing ages in subsequent years of the model. To allow for this variation, we advance the model forward by one year, using a closed-form analytic solution to the compartmental disease system differential equations that takes us from the start of the year to the end with constant rates over the time interval.

The solution that produces *S*_*t* + 1_ and *P*_*t* + 1_, given initial values *S*_*t*_ and *P*_*t*_, is

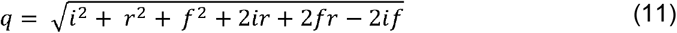

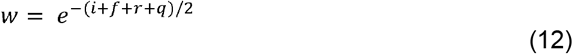

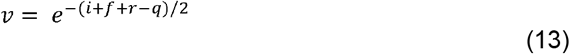

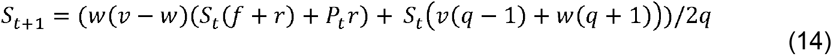

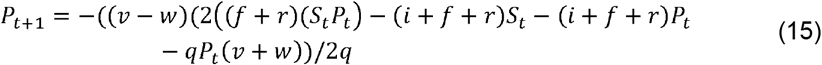

Model trajectories are produced by evaluating these equations 20 times to simulate 20 years, incrementing the age of each cohort each year. The resulting estimates of prevalence and deaths are then compared to the calibration targets, for each year by age group, for a given sex, as

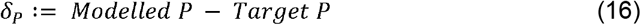

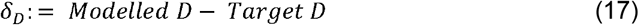

Note that *δ*_*P*_ and *δ*_*D*_ are each vectors indexed by age and year, and that their sign as well as magnitude are instrumental to the adjustments that occur. Technically, *i*_*b*_, *f*_*b*_, *r*_*b*_, *i*_*a*_, *f*_*a*_, and *r*_*a*_ are not adjusted directly, instead, we define the auxiliary parameters *P*_*b*_, *D*_*b*_, *P*_*b*_, and *D*_*b*_, which are age-indexed vectors that act on the underlying parameters element-wise. The adjusted rates are used to produce a model trajectory, which is used to calculate *δ*_*P*_ and *δ*_*D*_, which informs the adjustment of the auxiliary parameters. The adjusted rates are as follows.

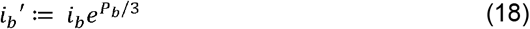

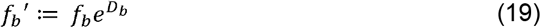

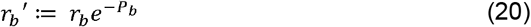

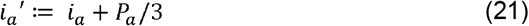

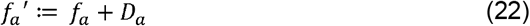

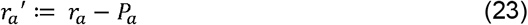

The auxiliary parameters *P*_*b*_, *D*_*b*_, *P*_*a*_, and are initialized to zero and subsequently adjusted to optimize the fit. This adjustment is weighted by year and is centred on a range of several age ‘filters’ that allow the optimization process to modify their behaviour at some ages without affecting others. The age filters are defined for ages spaced 15 years apart, starting at age zero and ending at age 105. We define *m*_*x*_, the filter at age *x*, to be the age-indexed vector which is zero up until age *x* − 15, increases linearly to a value of 1 at *x*, then linearly drops to 0 at age *x* +15. We also define *w*_*b*_ as a base year filter and *w*_*a*_ as a filter for all subsequent years (based on the annual percentage change), weighted towards earlier or later years, respectively. The year filters are used to make the annual percentage change parameters more sensitive to problems further into the future, and to make the base year parameters relatively less sensitive.

The auxiliary parameters are adjusted based on the value of *δ*_*P*_ and *δ*_*D*_. For example, *p*_*b*_ is modified as follows

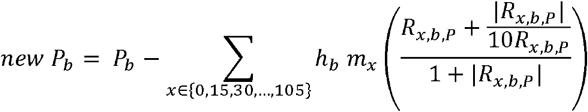

where *R*_*x,b*,*P*_ ≔ *δ*_*P*_ · (*m*_*x*_ =*z*_*b*_) and |*R*_*x,b*,*P*_ | is the element-wise absolute value. Note that the vector *m*_*x*_ is age-indexed while *w*_*b*_ is year-indexed, with the cross operation representing pairwise multiplication that is flattened into a vector indexed by age and year, which is appropriate for the dot product with *δ*_*P*_. The purpose of *R*_*x,b*,*P*_ is to reduce *P*_*b*_ around the ages where *δ*_*P*_ is too high and increase it where *δ*_*P*_ is too low, paying more attention to the early years, since *w*_*b*_ is weighted towards the start of the model. The effect is attenuated in subsequent iterations by the temperature parameter, *h*_*b*._ The modification of *D*_*b*_, *P*_*a*_, and *D*_*a*_ proceed in the same way, replacing the subscripts of *h* and *R* propriately. The algorithm switches back and forth between adjusting *P*_*b*_ and *D*_*b*_ in one iteration, then modifying *P*_*a*_ and *D*_*a*_ in the next.

The temperature parameters, *h*_*a*_ and *h*_*b*_, are reduced over subsequent iterations. The base year temperature, *h*_*b*_, starts higher than *h*_*a*_, but is reduced at a greater rate. This allows the conflict between the prevalence and mortality scores to slowly settle into a stable configuration. Incidence and remission are adjusted, in opposite directions, by the same underlying parameter; however, remission is modified at triple the rate (given that this parameter was inferred from the available input data).

The process runs over several hundred iterations, stopping when the previous 20 iterations produce scores (sum of the squares of each of the elements of *δ*_*P*_ and *δ*_*D*_, weighted by the WHO age standard (16)) all within 0.5% of each other. At this point the adjusted rates, as modified by the auxiliary parameters, are output as the final model parameters.

### Model output case studies

#### Part 1: historical model comparison

First, we show the output of our chronic disease generator model when using only a subset of the historical input data. We limited the GBD input data to 1990-2009, then ran the model from 2010-2019, allowing for an initial comparison of model performance against existing data. Figure 3 shows this comparison as a time series, for the top 20 chronic diseases in Australia (in terms of DALYs caused (14)). In this graph, the ‘rebased’ regression output, obtained by vertically aligning the regression trend to the 2019 value, is the calibration target of the simulation model. The initial model output is the result of the chronic disease model, using the initialization parameters of incidence, case fatality, and remission rates, forecast based on 1990-2009 data. The optimized model output is the result of calibrating the three rates to the prevalence (and mortality) target.

**Figure 3.**
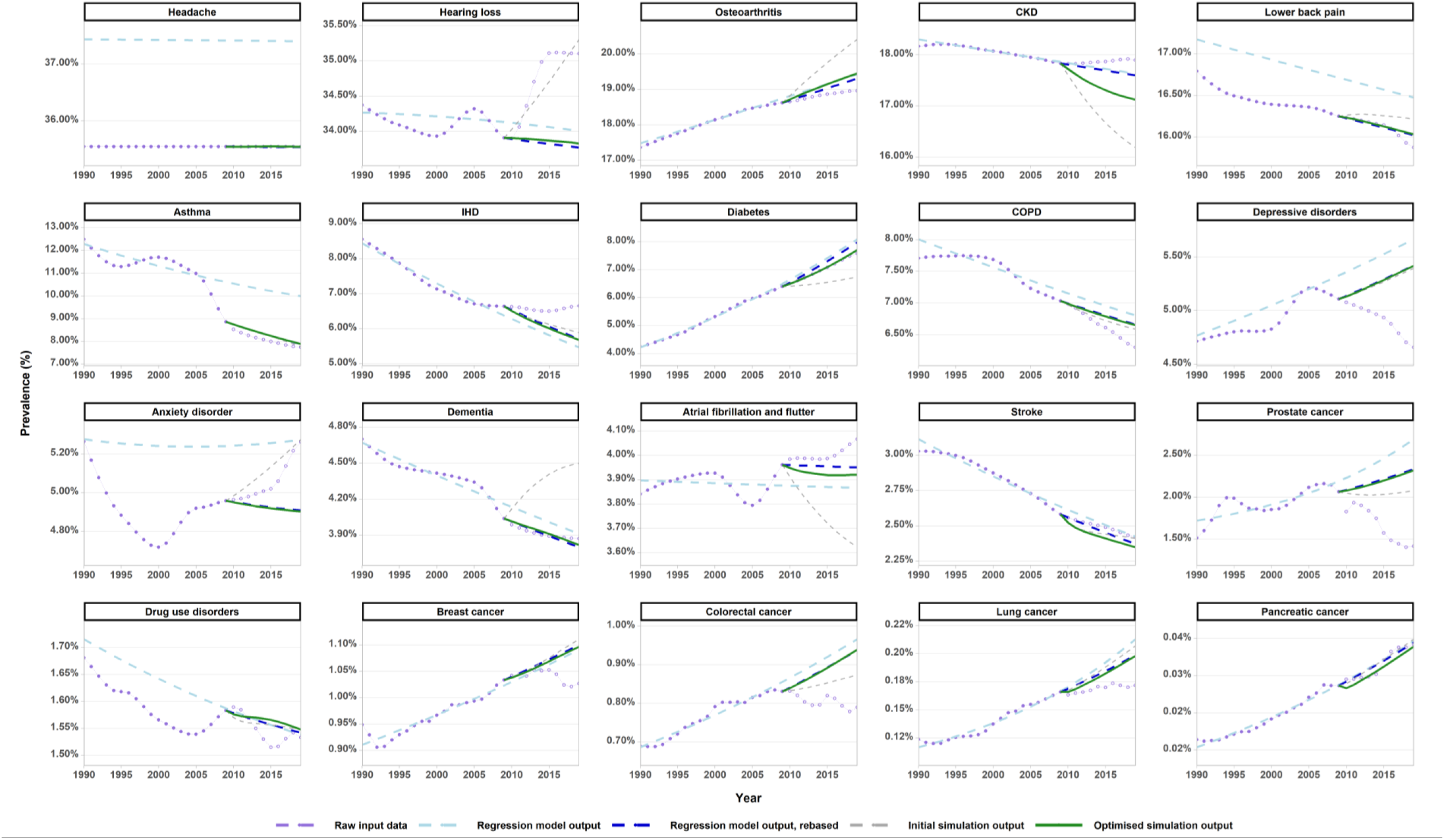
Historical model comparison - mean age-standardised prevalence for 20 diseases, 1990-2019. Standardised to the WHO-age-standard. (15) GBD input data not included in the regression & simulation models shown with open circles. ‘Rebased’ regression output, which shifts the start point of the regression line to the GBD 2019 datapoint, is the calibration target. Output for deaths shown in Supplementary Results.

Table 3 compares the fit of the resulting prevalence output (both from the initialization parameters regression output [step 2], and optimized model values [step 3]) in 2019 against GBD 2019 prevalence data.

**Table 3.**
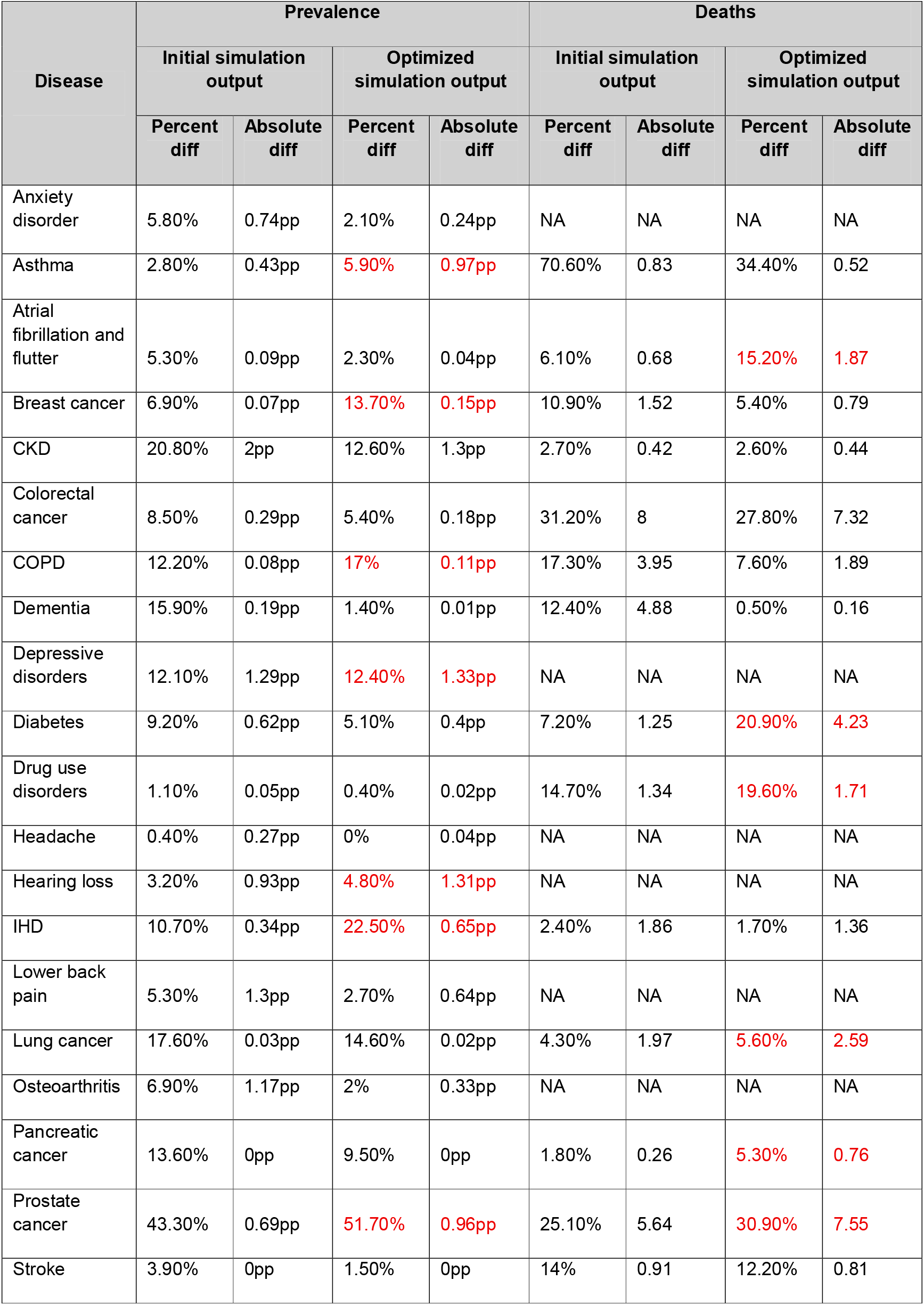
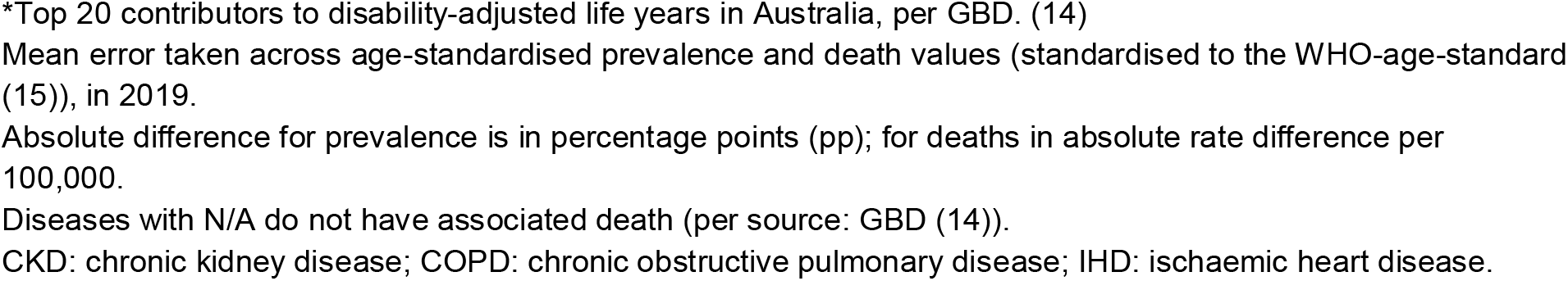
Comparison of subset model fit to GBD 2019 data, for top 20* chronic diseases in Australia.

| Disease | Prevalence |  |  |  | Deaths |  |  |  |
| --- | --- | --- | --- | --- | --- | --- | --- | --- |
|  | Initial simulation output |  | Optimized simulation output |  | Initial simulation output |  | Optimized simulation output |  |
|  | Percent diff | Absolute diff | Percent diff | Absolute diff | Percent diff | Absolute diff | Percent diff | Absolute diff |
| Anxiety disorder | 5.80% | 0.74pp | 2.10% | 0.24pp | NA | NA | NA | NA |
| Asthma | 2.80% | 0.43pp | 5.90% | 0.97pp | 70.60% | 0.83 | 34.40% | 0.52 |
| Atrial fibrillation and flutter | 5.30% | 0.09pp | 2.30% | 0.04pp | 6.10% | 0.68 | 15.20% | 1.87 |
| Breast cancer | 6.90% | 0.07pp | 13.70% | 0.15pp | 10.90% | 1.52 | 5.40% | 0.79 |
| CKD | 20.80% | 2pp | 12.60% | 1.3pp | 2.70% | 0.42 | 2.60% | 0.44 |
| Colorectal cancer | 8.50% | 0.29pp | 5.40% | 0.18pp | 31.20% | 8 | 27.80% | 7.32 |
| COPD | 12.20% | 0.08pp | 17% | 0.11pp | 17.30% | 3.95 | 7.60% | 1.89 |
| Dementia | 15.90% | 0.19pp | 1.40% | 0.01pp | 12.40% | 4.88 | 0.50% | 0.16 |
| Depressive disorders | 12.10% | 1.29pp | 12.40% | 1.33pp | NA | NA | NA | NA |
| Diabetes | 9.20% | 0.62pp | 5.10% | 0.4pp | 7.20% | 1.25 | 20.90% | 4.23 |
| Drug use disorders | 1.10% | 0.05pp | 0.40% | 0.02pp | 14.70% | 1.34 | 19.60% | 1.71 |
| Headache | 0.40% | 0.27pp | 0% | 0.04pp | NA | NA | NA | NA |
| Hearing loss | 3.20% | 0.93pp | 4.80% | 1.31pp | NA | NA | NA | NA |
| IHD | 10.70% | 0.34pp | 22.50% | 0.65pp | 2.40% | 1.86 | 1.70% | 1.36 |
| Lower back pain | 5.30% | 1.3pp | 2.70% | 0.64pp | NA | NA | NA | NA |
| Lung cancer | 17.60% | 0.03pp | 14.60% | 0.02pp | 4.30% | 1.97 | 5.60% | 2.59 |
| Osteoarthritis | 6.90% | 1.17pp | 2% | 0.33pp | NA | NA | NA | NA |
| Pancreatic cancer | 13.60% | 0pp | 9.50% | 0pp | 1.80% | 0.26 | 5.30% | 0.76 |
| Prostate cancer | 43.30% | 0.69pp | 51.70% | 0.96pp | 25.10% | 5.64 | 30.90% | 7.55 |
| Stroke | 3.90% | 0pp | 1.50% | 0pp | 14% | 0.91 | 12.20% | 0.81 |

\*Top 20 contributors to disability-adjusted life years in Australia, per GBD. (14)
Mean error taken across age-standardised prevalence and death values (standardised to the WHO-age-standard (15)), in 2019.
Absolute difference for prevalence is in percentage points (pp); for deaths in absolute rate difference per 100,000.
Diseases with N/A do not have associated death (per source: GBD (14)).
CKD: chronic kidney disease; COPD: chronic obstructive pulmonary disease; IHD: ischaemic heart disease.

The calibration step improved fit to 2019 GBD data across both targets for eight diseases, and across one target for 11 diseases. Prostate was the only disease with poorer fit across both outcomes – the trend for this disease in 2010-2019 changed direction in comparison to the data in the previous 20 year (Figure 3). This resulted in the prevalence projection, and hence the calibration target, did not closely match the data in 2019. Similar diverging patterns in forecast prevalence from 2010-2019 are seen for anxiety, depression, and hearing loss (Figure 3), and in forecast deaths for diabetes, atrial fibrillation and flutter, and to some extent lung cancer (Supplementary Figure 1).

#### Part 2: optimized model output

In Part 1 we demonstrated model outputs over a fixed hold-out period for a measure of predictive validity. Building on those results, we next use all available historical data to produce the final forecast model. The primary purpose of this model is to make ‘business-as-usual’ forecasts of coherent chronic disease parameters, for which a ground truth is not available to calibrate a model of the future to. Accordingly, the chronic disease generator model was run using the GBD 2021 dataset including 2010-2019 data, (2020-2021 data excluded, and GBD 2023 not publicly available at the time of analysis).

On average, the optimization reached the stopping criterion after 380 iterations (details in Supplementary Material).

Figure 4 presents 20-year prevalence forecasts for each of the same diseases shown in Part 1. The difference between the target in 2039 and both a) the initial unoptimized output produced from the initialization parameters, and b) the final optimized values produced from calibrated rates, are summarised in Table 4. Unlike in Part 1, which compared model outputs to raw data, this comparison estimates the difference to the forecast prevalence and mortality rate targets from Step 2. The calibration model improved fit to the targets for all outcomes.

**Table 4.**
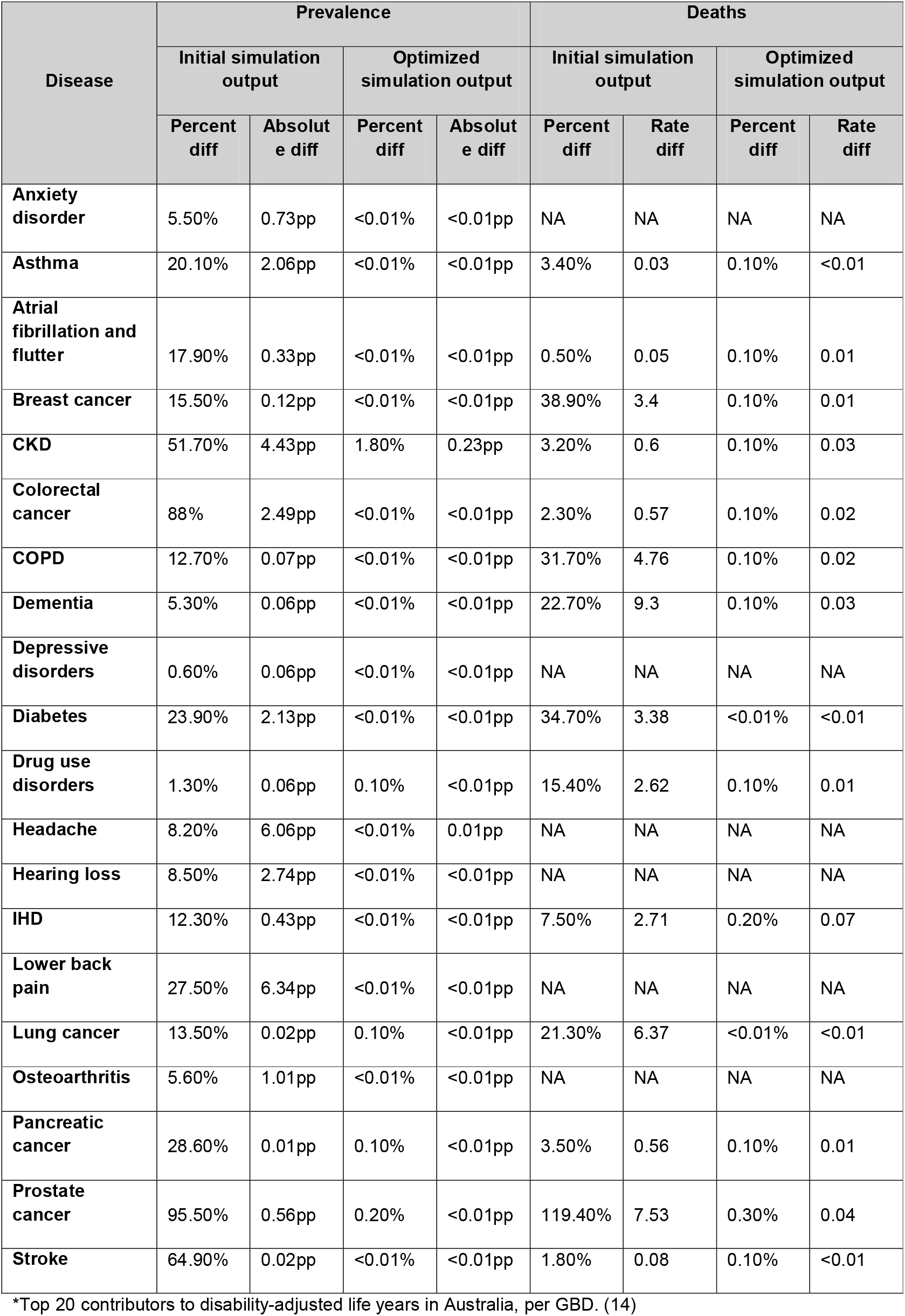
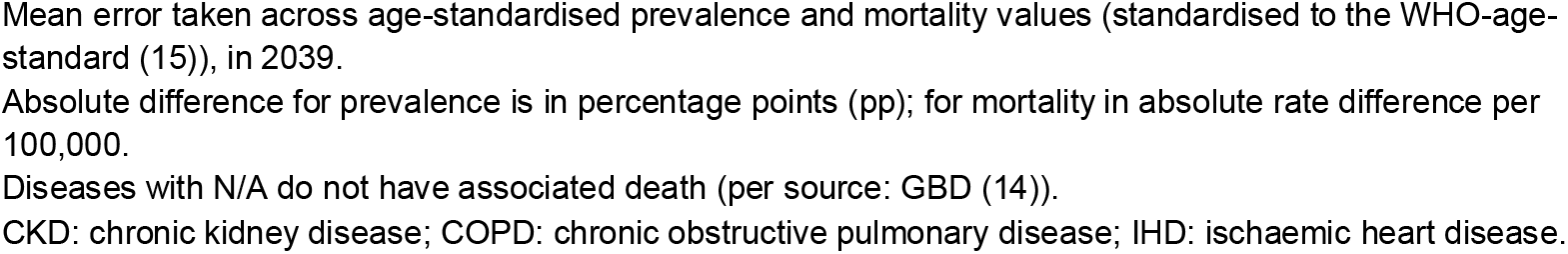
Comparison of model fit calibration targets in 2019, for top 20* chronic diseases in Australia.

| Disease | Prevalence |  |  |  | Deaths |  |  |  |
| --- | --- | --- | --- | --- | --- | --- | --- | --- |
|  | Initial simulation output |  | Optimized simulation output |  | Initial simulation output |  | Optimized simulation output |  |
|  | Percent diff | Absolute diff | Percent diff | Absolute diff | Percent diff | Rate diff | Percent diff | Rate diff |
| **Anxiety disorder** | 5.50% | 0.73pp | <0.01% | <0.01pp | NA | NA | NA | NA |
| **Asthma** | 20.10% | 2.06pp | <0.01% | <0.01pp | 3.40% | 0.03 | 0.10% | <0.01 |
| **Atrial fibrillation and flutter** | 17.90% | 0.33pp | <0.01% | <0.01pp | 0.50% | 0.05 | 0.10% | 0.01 |
| **Breast cancer** | 15.50% | 0.12pp | <0.01% | <0.01pp | 38.90% | 3.4 | 0.10% | 0.01 |
| **CKD** | 51.70% | 4.43pp | 1.80% | 0.23pp | 3.20% | 0.6 | 0.10% | 0.03 |
| **Colorectal cancer** | 88% | 2.49pp | <0.01% | <0.01pp | 2.30% | 0.57 | 0.10% | 0.02 |
| **COPD** | 12.70% | 0.07pp | <0.01% | <0.01pp | 31.70% | 4.76 | 0.10% | 0.02 |
| **Dementia** | 5.30% | 0.06pp | <0.01% | <0.01pp | 22.70% | 9.3 | 0.10% | 0.03 |
| **Depressive disorders** | 0.60% | 0.06pp | <0.01% | <0.01pp | NA | NA | NA | NA |
| **Diabetes** | 23.90% | 2.13pp | <0.01% | <0.01pp | 34.70% | 3.38 | <0.01% | <0.01 |
| **Drug use disorders** | 1.30% | 0.06pp | 0.10% | <0.01pp | 15.40% | 2.62 | 0.10% | 0.01 |
| **Headache** | 8.20% | 6.06pp | <0.01% | 0.01pp | NA | NA | NA | NA |
| **Hearing loss** | 8.50% | 2.74pp | <0.01% | <0.01pp | NA | NA | NA | NA |
| **IHD** | 12.30% | 0.43pp | <0.01% | <0.01pp | 7.50% | 2.71 | 0.20% | 0.07 |
| **Lower back pain** | 27.50% | 6.34pp | <0.01% | <0.01pp | NA | NA | NA | NA |
| **Lung cancer** | 13.50% | 0.02pp | 0.10% | <0.01pp | 21.30% | 6.37 | <0.01% | <0.01 |
| **Osteoarthritis** | 5.60% | 1.01pp | <0.01% | <0.01pp | NA | NA | NA | NA |
| **Pancreatic cancer** | 28.60% | 0.01pp | 0.10% | <0.01pp | 3.50% | 0.56 | 0.10% | 0.01 |
| **Prostate cancer** | 95.50% | 0.56pp | 0.20% | <0.01pp | 119.40% | 7.53 | 0.30% | 0.04 |
| **Stroke** | 64.90% | 0.02pp | <0.01% | <0.01pp | 1.80% | 0.08 | 0.10% | <0.01 |

\*Top 20 contributors to disability-adjusted life years in Australia, per GBD. (14)
Mean error taken across age-standardised prevalence and mortality values (standardised to the WHO-age-standard (15)), in 2039.
Absolute difference for prevalence is in percentage points (pp); for mortality in absolute rate difference per 100,000.
Diseases with N/A do not have associated death (per source: GBD (14)).
CKD: chronic kidney disease; COPD: chronic obstructive pulmonary disease; IHD: ischaemic heart disease.

**Figure 4.**
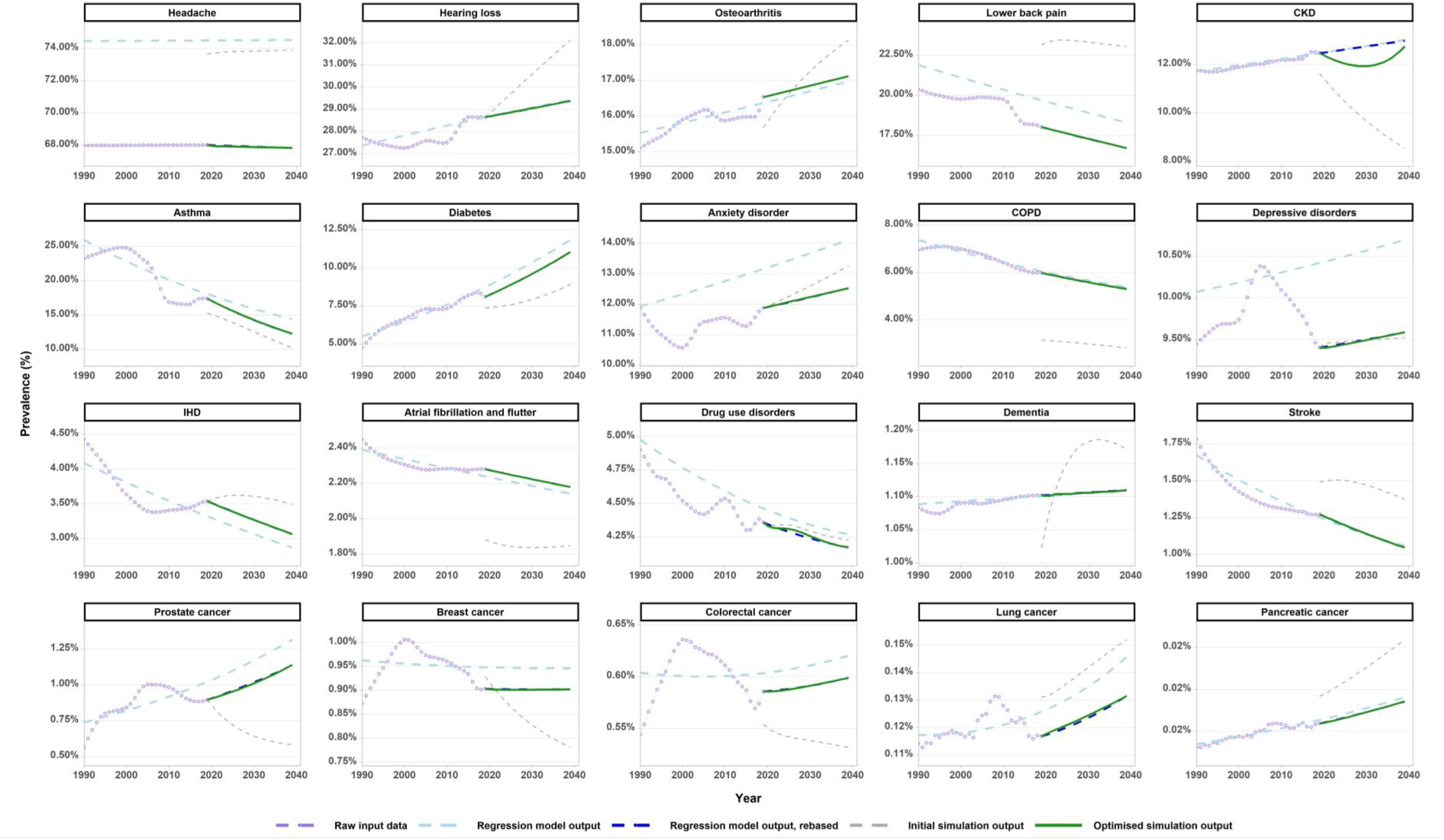
Full model comparison - mean age-standardised prevalence for 20 diseases, 2019-2039. Standardised to the WHO-age-standard.(15) Rebased regression output has been scaled to GBD 2019 raw input data as a start point. This is the calibration target for the simulation model.

## Discussion

The chronic disease rate generator presented in this paper is a novel, data driven approach to producing forecasts for a compartmental disease model, for any (chronic) outcome and in any setting. The model generates internally consistent trajectories of prevalence, incidence, remission, and case fatality using routinely available data. The full-data model resulted in good fit to calibration targets (Part 2 results), being prevalence and death forecasts, for each of 20 tested diseases, through changes to initial projections of disease incidence, case fatality, and remission rates.

This method solves two key issues when obtaining disease data for modelling purposes, of 1) the availability of current/historic disease data, and 2) the lack of coherence that results from projecting rates for a given disease independently. The latter was illustrated by the difference between directly forecast prevalence values and prevalence derived from unoptimized disease rate forecasts (Figures 3 & 4). The model requires minimal manual handling with its data-driven approach and is therefore suited for modelling of multiple diseases simultaneously. It can also be routinely updated with future iterations of the GBD (or other data sources). Further, the model can be easily adapted for acute disease forecasting (code provided in GitHub).

These forecasts are primed for use in simulation modelling, providing an important baseline for modelling interventions targeting diseases, either directly (e.g., via a new health technology that reduces the case fatality rate for a given disease) or upstream via disease risk factors (e.g., interventions targeting smoking, which is linked to 30+ diseases (17)). Such information is crucial for informing policy.

There are limitations to this method. While the historical data subset model (Part 1 results; excluding data from 2010-2019) was used to test the model’s predictive validity, (18) the model was not externally validated due to a lack of externally produced disease prevalence forecasts available for comparison. The approach taken assumes that disease trends follow the trends seen over the past 30 years. This implicitly incorporates the effect of public health policies occurring over that period and assumes that the momentum of these actions in healthcare will continue. This is just one statistical approach to forecasting ‘business-as-usual’ disease trends, and further out projections become increasingly uncertain.

It is important to note that while the disease generator produces coherent projections for each given disease, there is not internal coherence between individual disease projections and all-cause morbidity and mortality for a given population. In other words, the mortality rate summed across all diseases would not necessarily be that of a comparable forecast of all-cause mortality rates. If one has a view that a separate all-cause mortality rate forecast is the ‘truth’ one is working to, then disease specific rates could be scaled to achieve this external calibration target.

## Data Availability

The dataset used in the current study are available from the IHME website: https://vizhub.healthdata.org/gbd-results/
The model used to generate results in the current study are available from GitHub: https://github.com/population-interventions/CompartmentalDiseaseGenerator

https://github.com/population-interventions/CompartmentalDiseaseGenerator

## Abbreviations

CKD: chronic kidney disease
COPD: chronic obstructive pulmonary disease
DALY: disability-adjusted life year
GBD: Global Burden of Disease
GOF: Goodness of Fit
IHD: ischaemic heart disease
WHO: World Health Organisation

## Declarations

### Ethics approval and consent to participate

Ethics approval was not required for this research, as only publicly available, deidentified data was used.

### Consent for publication

Not applicable

### Availability of data and materials

The dataset used in the current study are available from the IHME website: https://vizhub.healthdata.org/gbd-results/

The model used to generate results in the current study are available from GitHub: https://github.com/population-interventions/CompartmentalDiseaseGenerator

### Authors’ contributions

T.W. and T.B. conceptualised the model. T.W. produced the model code. S.H. and T.W. wrote the manuscript. S.H. prepared the graphs. All authors contributed to editing the draft and approving the final manuscript.

## Acknowledgements

Not applicable

