## Supplementary Data for "Specifying prospective compartmental models of chronic disease"

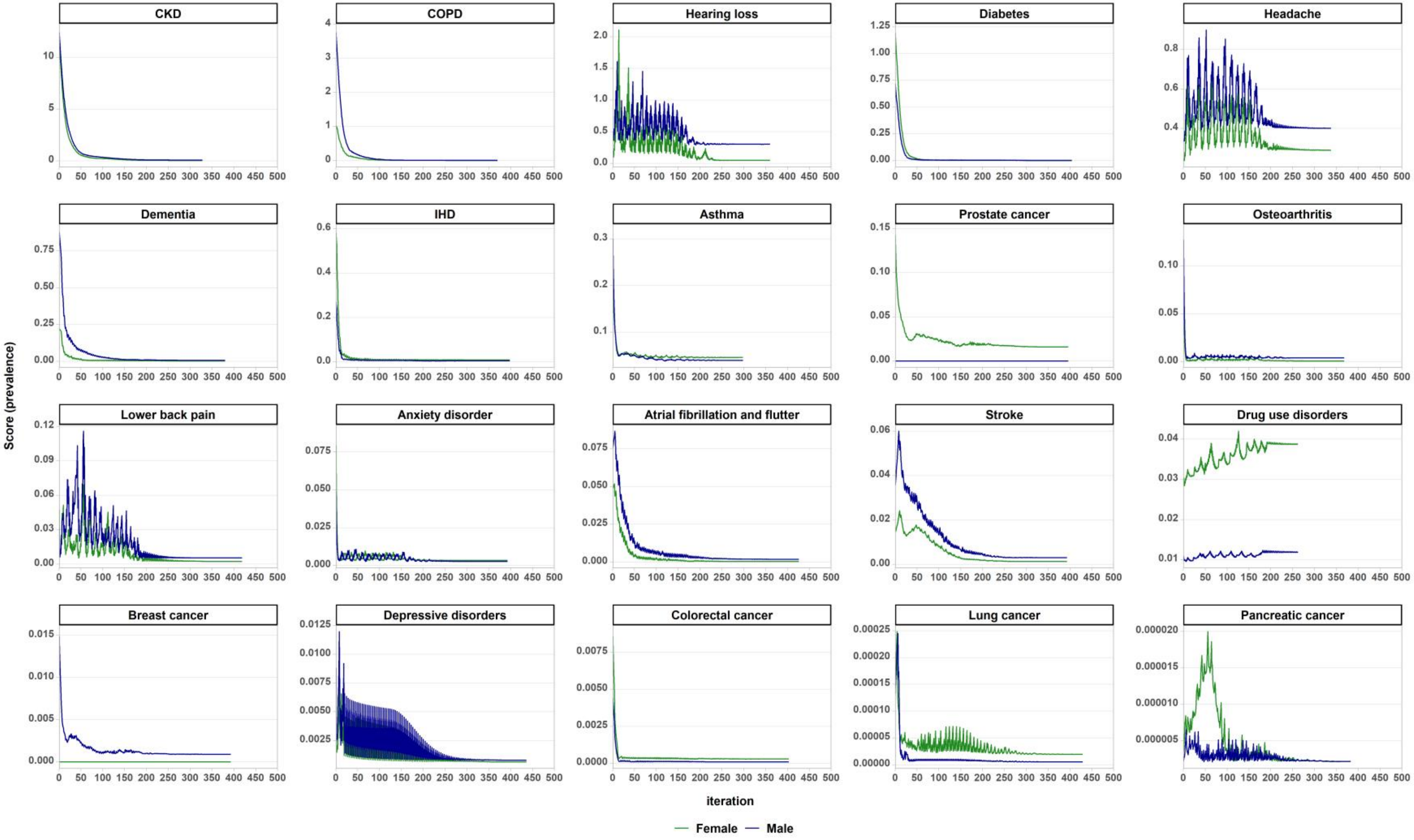


**Supplementary Figure 1. Calibration model process: scores for prevalence target by sex.**

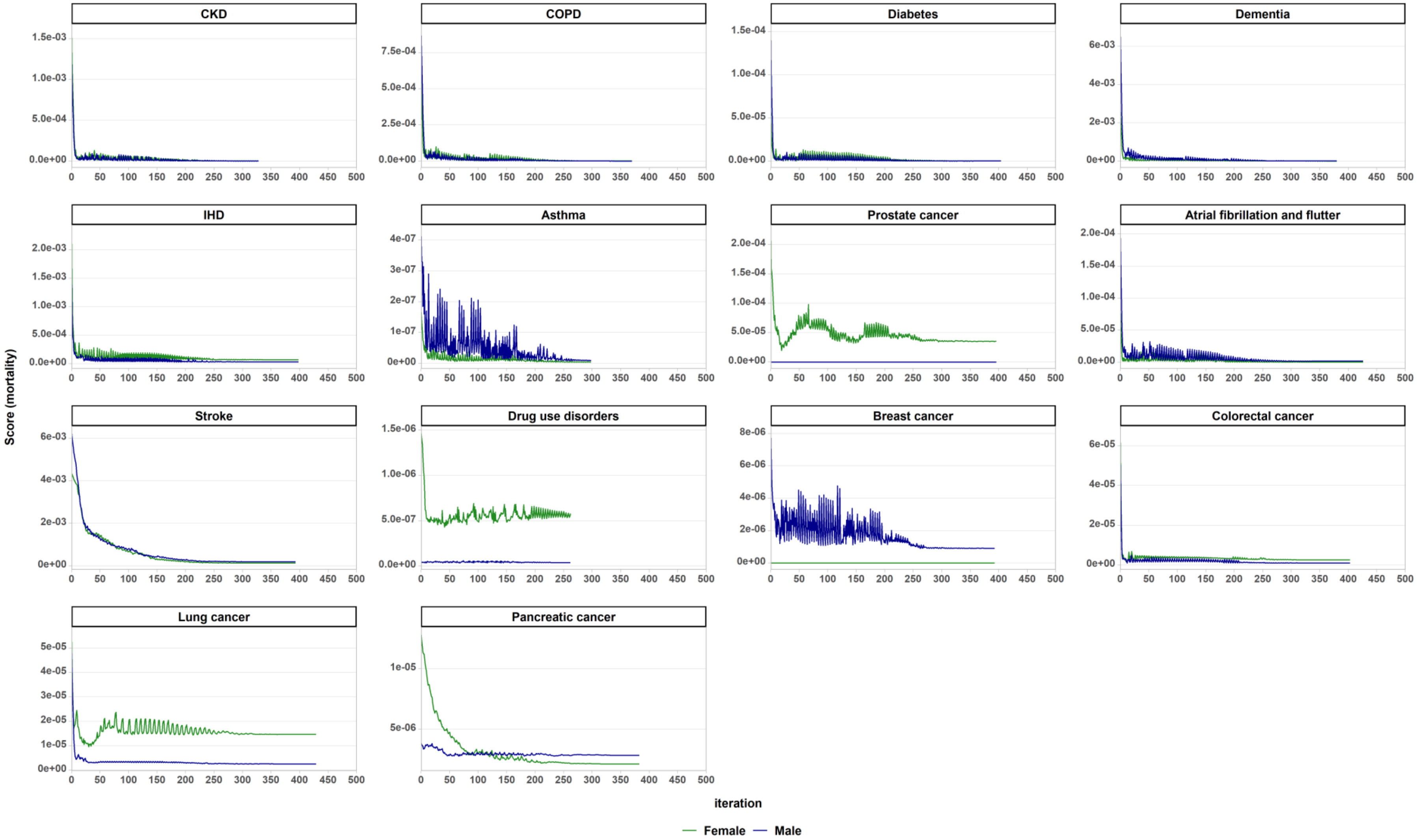


**Supplementary Figure 2. Calibration model process: scores for mortality rate target by sex.**

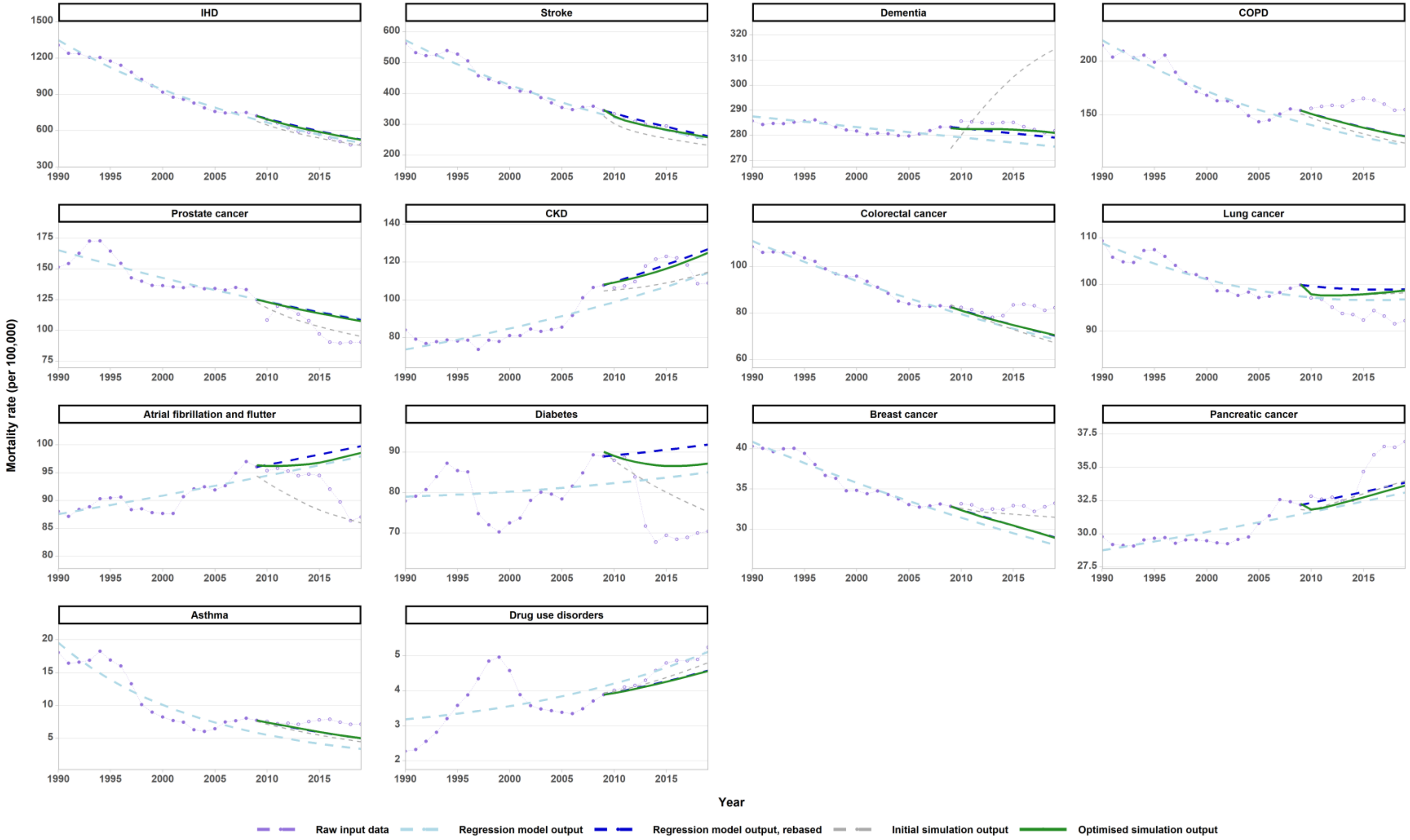


**Supplementary Figure 3. Historical model comparison - mean age-standardised mortality rates, 1990-2019.** GBD input data not included in the regression & simulation models shown with open circles. 'Rebased' regression output, which shifts the start point of the regression line to the GBD 2019 datapoint, is the calibration target. Mortality rates standardised using WHO-age-standard.(1)

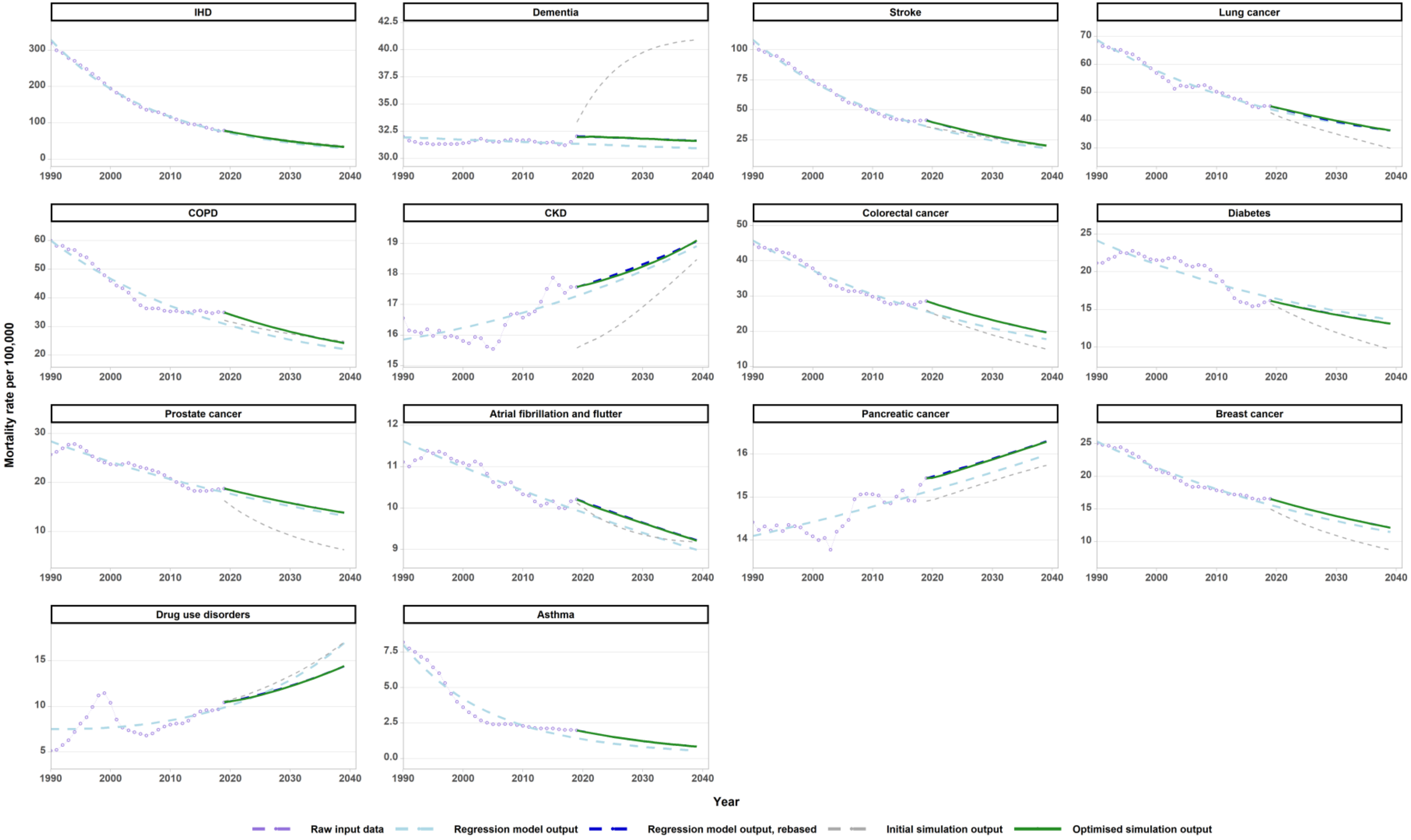


**Supplementary Figure 4. Full model comparison - mean age-standardised mortality rates for 14 diseases, 2019-2039.** Diseases with no associated deaths (as per GBD) not included. Mortality rates standardised using WHO-age-standard.(1)
